# Effectiveness of Non-Pharmaceutical Interventions on Child and Staff COVID-19 Cases in US Summer Camps

**DOI:** 10.1101/2021.02.18.21250271

**Authors:** Helen H. Suh, Julianne Meehan, Laura Blaisdell, Laurie Browne

**Affiliations:** Department of Civil & Environmental Engineering, Tufts University, Medford, MA; Environmental Health & Engineering, Inc., Newton, MA; Department of Pediatrics, Maine Medical Center, Portland, ME & Camp Winnebago, Fayette, ME; American Camp Association, Martinsville, IN

**Author notes:** **Corresponding Author:** Helen H. Suh, Department of Civil & Environmental Engineering, Tufts University, Medford, MA 02155. **Role of Funder:** ACA provided survey responses, but had no role in the analysis of study data; Dr. Browne reviewed the manuscript. **Contributors Statement Page:** Dr. Suh conceptualized and designed the study, drafted, reviewed and revised the manuscript; Ms. Meehan conducted data analyses and reviewed the manuscript; Dr. Blaisdell critically reviewed the manuscript; Dr. Browne designed the data collection instruments, collected data, and reviewed the manuscript; All authors approved the final manuscript as submitted and agree to be accountable for all aspects of the work.

## Abstract

**Background:** Most camps remained closed during Summer 2020, due to concerns regarding child transmission of SARS-CoV-2 and limited information about the effectiveness of non-pharmaceutical interventions (NPIs) within child congregate settings.

**Methods:** We surveyed US camps about on-site operations, camper and staff demographics, COVID-19 cases amongst campers and staff, and NPI usage as related to pre-camp quarantines, facial coverings, physical distancing, cleaning, and facility modifications. For all NPIs, save quarantines, responses were provided on a 5-point Likert scale format.

**Results:** Within 486 on-site camps, a range of NPIs were instituted, most often related to reduced camper interactions, staff face coverings, cleaning, and hand hygiene. Camper facial coverings were less common, with campers always wearing masks at ∼34% of the camps. Approximately 15% of camps reported 1+ confirmed COVID-19 case in either campers or staff, with three camps reporting a COVID outbreak. In both single and multi-NPI analyses, the risk of COVID-19 cases was lowest when campers always wore facial coverings. While less effective, constant use of staff facial coverings and targeted physical distancing measures, but not pre-camp quarantine, also reduced COVID-19 risks.

**Conclusions:** We found constant facial coverings, especially for campers, and targeted physical distancing measures to reduce risks of SARS-CoV-2 transmission within summer camps. Our findings provide valuable guidance for future operations of camp and other child congregate settings with regard to efficient and effective NPI usage to mitigate SARS-CoV-2 infection.

**What’s Known on This Subject:** Approximately 82% of US overnight camps did not open during Summer 2020 due to concerns regarding children’s ability to transmit SARS-CoV-2. Camps that did operate during this time instituted varied non-pharmaceutical interventions (NPIs) to reduce SARS-CoV-2 transmission, with little information available on the effectiveness of these NPIs within child congregate settings. Large population-based studies are needed to improve our understanding of the extent of SARS-CoV-2 infection amongst children and their caregivers and to determine whether and to what degree child congregate programs can safely open during the pandemic.

**What This Study Adds:** Our study, the largest survey of COVID-19 cases in child congregate settings at the national level, provides new information on the relative effectiveness of NPIs on mitigating COVID cases among children and staff within camp settings. We showed COVID-19 case rates in campers and staff to be low relative to corresponding case rates in the US and found constant camper facial coverings to be the most effective risk reduction method for SARS-CoV-2 transmission within camps. While less effective, constant use of staff facial coverings and targeted physical distancing measures, but not pre-camp quarantines, were also shown to reduce COVID-19 risks. Our findings has important implications for child congregate settings, helping to guide their successful opening and operation.

## Introduction

Each year, summer camps in the United States (US) host more than 26 million children and employ 1.5 million staff of all ages, race/ethnicities, genders, and socio-economic position [1]. Alongside the closing of schools, camps were massively disrupted across the US as a result of the coronavirus disease 2019 (COVID-19) pandemic. Many camp programs, including approximately 82% of US overnight camps [1], did not open during Summer 2020, due to a lack of understanding of (1) the degree to which SARS-CoV-2 is transmitted in within child congregate settings and (2) the non-pharmaceutical interventions (NPIs) needed to minimize this transmission.

To date, few studies have been conducted that examine SARS-CoV-2 transmission and NPI effectiveness in camp settings, with evidence to date anecdotal in nature or based on small sample sizes. This evidence shows that camps that operated during Summer 2020 experienced varying degrees of success in mitigating SARS-CoV-2 transmission, possibly due to differences in the use of non-pharmaceutical interventions (NPI). SARS-CoV-2 transmission, for example, was low within four Maine overnight camps that implemented prearrival quarantine, pre- and post-arrival testing and symptom screening, cohorting, face covering use, physical distancing, enhanced hygiene measures, cleaning and disinfecting, and maximal outdoor programming [2]. In contrast, an overnight camp in Georgia experienced significant SARS-CoV-2 transmission among campers and staff [3], with an overall attack rate of 44%. Attack rates increased with length of time spent at the camp. Transmission was attributed to large number of campers sleeping in the same cabin and frequent singing and cheering without the regular use of facial coverings. Similarly, an outbreak of COVID-19 occurred at a boys’ overnight summer school retreat in Wisconsin [4]. Among 152 attendees, 116 (76%) were classified as having confirmed or probable COVID-19. At this retreat, organizers required documentation of a negative prearrival RT-PCR result, 7-day prearrival quarantine, and outdoor programming, but did not implement other recommended NPIs.

In this paper, we use data from a nationwide survey of camps operating during Summer 2020 to estimate the prevalence of COVID-19 cases amongst campers and staff and its relation to individual and multiple NPIs instituted at these camps.

## Methods

The study was approved by the institutional review board at Tufts University.

As part of the American Camp Association (ACA)’s annual survey of US camps, we asked camps that operated in-person programs in summer 2020 about camper and staff demographics, COVID-19 cases, and non-pharmaceutical interventions (NPI). ACA’s online survey was distributed in September 2020 via weekly e-newsletter, direct email, and camp networks. Respondents answered anonymously on behalf of a single camp or multiple camps (if the respondent operated multiple sites). Camps were asked to provide information about (1) their state of operation, (2) the state and countries in which their campers resided, and (3) the number and demographic characteristics (as percent) of campers and staff, with single site camps providing answers by week and multi-site camps as aggregate information over the summer.

COVID-19-related questions included those related to the number of confirmed and suspected COVID-19 cases for campers and staff, again reported by week by single site camps and as aggregate information by multi-site camps for each of their camps. NPI-related questions included those about camp policies and NPI usage with regard to camper and staff quarantines at home or at camp, staff and camper facial coverings, reduced capacity, activity cohorts, physical distancing, hand sanitizing, increased cleaning, decreased visitors, modified programs, and modified sleeping, dining, and bathroom arrangements. For all questions about NPIs, save those about quarantines, responses were provided on a 5-point Likert scale format, as always, often, sometimes, rarely and never.

Camp respondents leaving >50% of overall responses blank were omitted from data analysis. Data were analyzed using descriptive statistics, including summary statistics and graphical analysis of the time trends in COVID-19 cases and their relation to regional case rates. NPI effectiveness alone and in pairs was examined using risk ratios, with analyses of multiple NPIs performed based on total cases for specific NPI combinations. These combinations were selected based on their observed effectiveness when examined alone and on their combined use among many camps. All analyses were performed using R statistical software (R version 4.0.3).

## Results

A total of 1,193 single and multi-site camps completed the survey, representing 1,489 camps, as shown in Table 1. Camps were located in 49 states and the District of Columbia, with 26% located in the Midwest, 23% in the Northeast, 22% in the South, and 16% in the West. The racial and socioeconomic distribution of campers mirrored that of the entire camp population [1]. Approximately 73% of campers identified as White, 8.3% Black, 6.0% Hispanic, 4.2% Asian, and 6.0% bi- or multi-racial, while about 48% of campers were from middle income, 34% from high, and 18.5% from low income households.

**Table 1.** Summary of the Number of Camps, Campers and Confirmed COVID-19 Cases: Summer 2020

| Camp Descriptor | Number<sup>a</sup> |  | Confirmed Cases<sup>b</sup> |  |  |  |
| --- | --- | --- | --- | --- | --- | --- |
|  | Camps | Children | Camps | Camper | Staff | Total |
| **Operations for Summer 2020** |  |  |  |  |  |  |
| *All surveyed camps* | 1489 | 89,805 | 74 | 30 | 72 | 102 |
| *Onsite* | 486 | 89,635 | 74 | 30 | 72 | 102 |
| **Camp Type (Onsite Only)** |  |  |  |  |  |  |
| *Overnight* | 59 | 16,228 | 10 | 11 | 30 | 41 |
| *Day* | 206 | 25,826 | 52 | 16 | 29 | 45 |
| *Combination* | 220 | 47,581 | 12 | 3 | 13 | 16 |
| *Missing* | 1 | 0 | 0 | 0 | 0 | 0 |
| **Camp Type (Onsite Only)** |  |  |  |  |  |  |
| *Single camp* | 295 | 70,982 | 36 | 27 | 65 | 92 |
| *Multi-site camp* | 191 | 18,653 | 38 | 3 | 7 | 10 |
| **Region (Onsite Only)** |  |  |  |  |  |  |
| *Midwest* | 124 | 17,259 | 34 | 10 | 30 | 40 |
| *Northeast* | 113 | 24,665 | 8 | 5 | 12 | 17 |
| *South* | 108 | 20,605 | 15 | 9 | 18 | 27 |
| *West* | 78 | 11,696 | 10 | 1 | 7 | 8 |
| *Missing* | 63 | 15,410 | 5 | 5 | 5 | 10 |

<sup>a</sup> Number camps that responded to the survey; number of children onsite for summer 2020 estimated as unique campers
<sup>b</sup> Number of camps that reported cases; number of confirmed camper and staff cases

486 of the responding camps, serving about 90,000 campers, operated on-site during summer 2020, including 59 overnight, 206 day, and 220 combination day, overnight, and rental camps (with 1 missing response). Day and overnight single site camps operated in all regions of the US, with lower rates of operation in the West and highest rates of operation in the Northeast (Figure 1). Rates of operation for the single-site camps were highest during the middle of the summer in all regions of the US, as shown by the mid-summer peak in the number of camps, children, and staff at these single-site camps.

**Figure 1.**
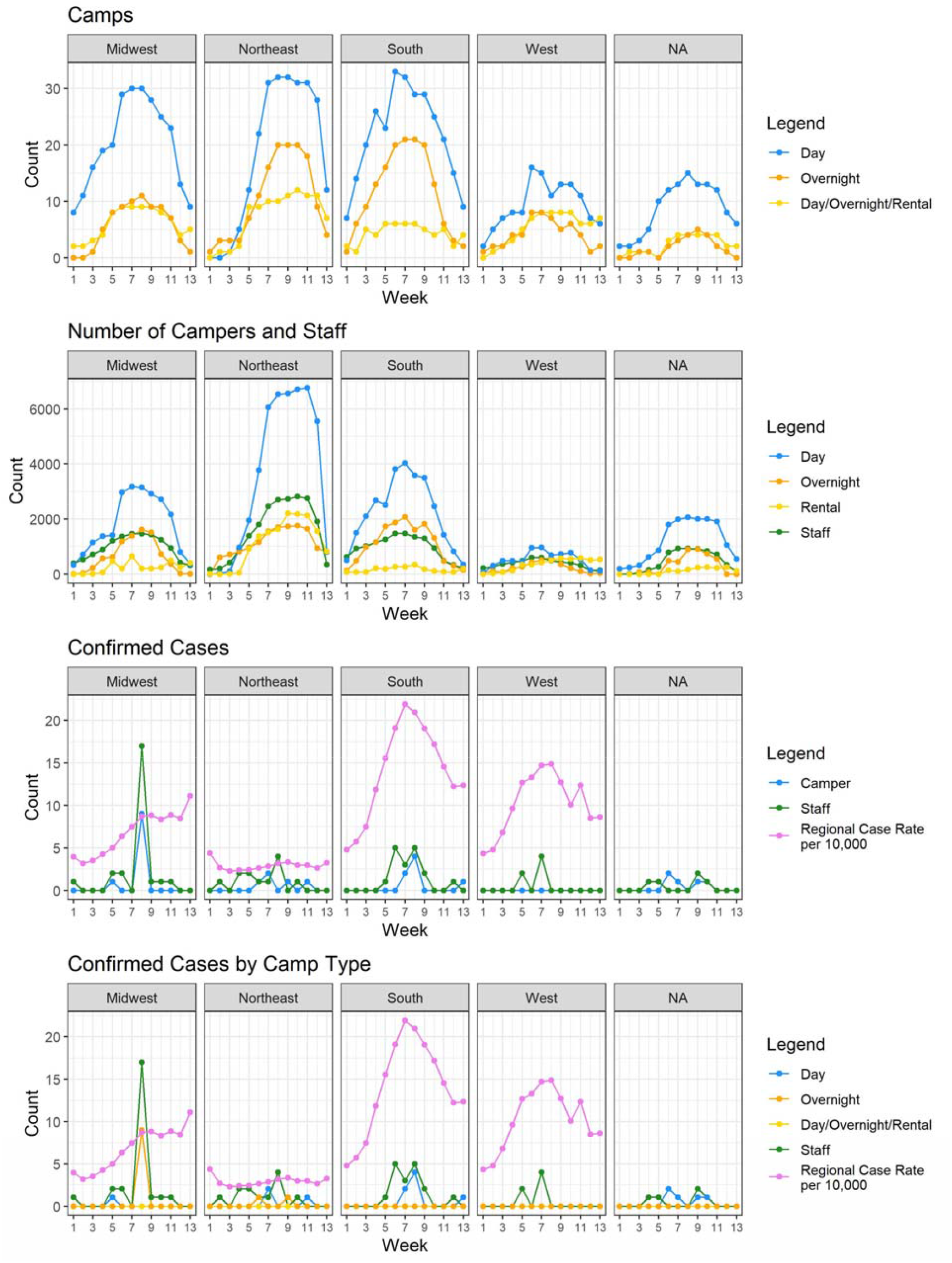
Weekly number of single site camps and associated campers, staff, and cases: by camp type, region.

NPI usage and COVID-19 cases across on-site camps are shown in Table 2. Most camps reported constant use of NPIs related to staff facial coverings (69%), reduced capacity (89%), smaller cohort sizes (86%), increased cleaning (95%), and more frequent handwashing (96%). Constant physical distancing (66%) and face coverings among campers (33%) was reported by a smaller number of camps. Correspondingly, approximately one-third of camps required campers and staff to quarantine at home prior to attending camp, with fewer camps requiring staff to quarantine at camp and only 18 requiring campers to quarantine at camp. Modifications to sleeping arrangements were fairly typical amongst overnight and combination overnight/day/rental camps, while altered dining and bathroom arrangements were more common at day and combination camps.

**Table 2.**
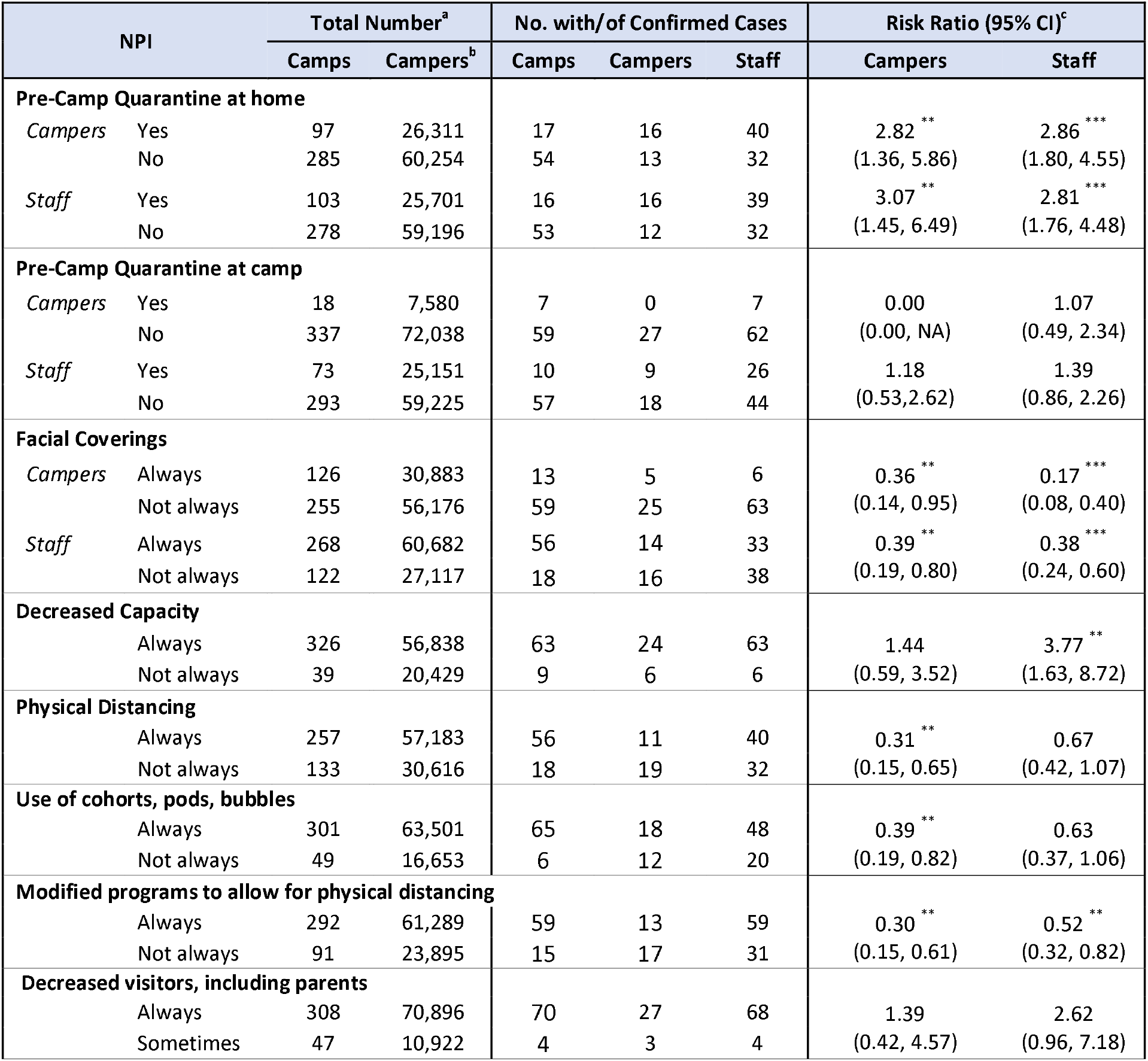
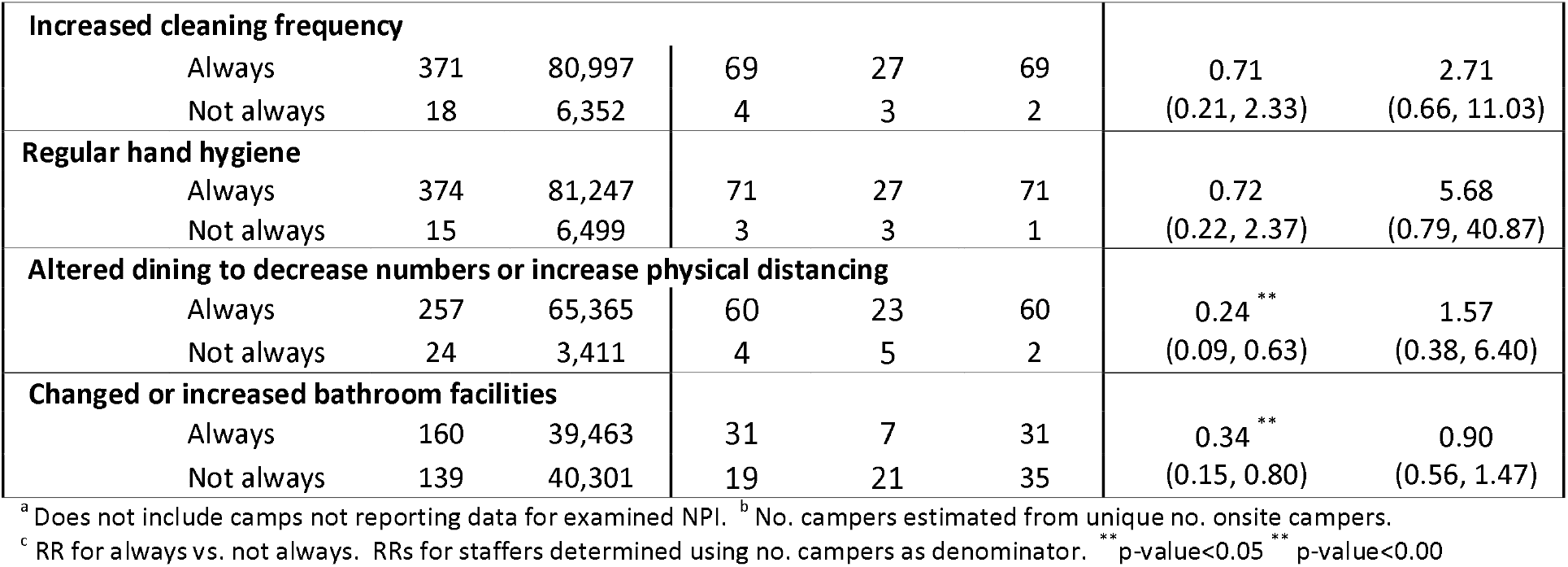
NPI Usage and Camper and Staff Number and COVID-19 Cases

| Table 2. NPI Usage and Camper and Staff Number and COVID-19 Cases |  |  |  |  |  |  |  |  |
| --- | --- | --- | --- | --- | --- | --- | --- | --- |
| NPI |  | Total Number<sup>a</sup> |  | No. with/of Confirmed Cases |  |  | Risk Ratio (95% CI)<sup>c</sup> |  |
|  |  | Camps | Campers<sup>b</sup> | Camps | Campers | Staff | Campers | Staff |
| Pre-Camp Quarantine at home |  |  |  |  |  |  |  |  |
| Campers | Yes | 97 | 26,311 | 17 | 16 | 40 | 2.82 ** | 2.86 *** |
|  | No | 285 | 60,254 | 54 | 13 | 32 | (1.36, 5.86) | (1.80, 4.55) |
| Staff | Yes | 103 | 25,701 | 16 | 16 | 39 | 3.07 ** | 2.81 *** |
|  | No | 278 | 59,196 | 53 | 12 | 32 | (1.45, 6.49) | (1.76, 4.48) |
| Pre-Camp Quarantine at camp |  |  |  |  |  |  |  |  |
| Campers | Yes | 18 | 7,580 | 7 | 0 | 7 | 0.00 | 1.07 |
|  | No | 337 | 72,038 | 59 | 27 | 62 | (0.00, NA) | (0.49, 2.34) |
| Staff | Yes | 73 | 25,151 | 10 | 9 | 26 | 1.18 | 1.39 |
|  | No | 293 | 59,225 | 57 | 18 | 44 | (0.53, 2.62) | (0.86, 2.26) |
| Facial Coverings |  |  |  |  |  |  |  |  |
| Campers | Always | 126 | 30,883 | 13 | 5 | 6 | 0.36 ** | 0.17 *** |
|  | Not always | 255 | 56,176 | 59 | 25 | 63 | (0.14, 0.95) | (0.08, 0.40) |
| Staff | Always | 268 | 60,682 | 56 | 14 | 33 | 0.39 ** | 0.38 *** |
|  | Not always | 122 | 27,117 | 18 | 16 | 38 | (0.19, 0.80) | (0.24, 0.60) |
| Decreased Capacity |  |  |  |  |  |  |  |  |
|  | Always | 326 | 56,838 | 63 | 24 | 63 | 1.44 | 3.77 ** |
|  | Not always | 39 | 20,429 | 9 | 6 | 6 | (0.59, 3.52) | (1.63, 8.72) |
| Physical Distancing |  |  |  |  |  |  |  |  |
|  | Always | 257 | 57,183 | 56 | 11 | 40 | 0.31 ** | 0.67 |
|  | Not always | 133 | 30,616 | 18 | 19 | 32 | (0.15, 0.65) | (0.42, 1.07) |
| Use of cohorts, pods, bubbles |  |  |  |  |  |  |  |  |
|  | Always | 301 | 63,501 | 65 | 18 | 48 | 0.39 ** | 0.63 |
|  | Not always | 49 | 16,653 | 6 | 12 | 20 | (0.19, 0.82) | (0.37, 1.06) |
| Modified programs to allow for physical distancing |  |  |  |  |  |  |  |  |
|  | Always | 292 | 61,289 | 59 | 13 | 59 | 0.30 ** | 0.52 ** |
|  | Not always | 91 | 23,895 | 15 | 17 | 31 | (0.15, 0.61) | (0.32, 0.82) |
| Decreased visitors, including parents |  |  |  |  |  |  |  |  |
|  | Always | 308 | 70,896 | 70 | 27 | 68 | 1.39 | 2.62 |
|  | Sometimes | 47 | 10,922 | 4 | 3 | 4 | (0.42, 4.57) | (0.96, 7.18) |

|  |  |  |  |  |  |  |  |
| --- | --- | --- | --- | --- | --- | --- | --- |
| **Increased cleaning frequency** |  |  |  |  |  |  |  |
| Always | 371 | 80,997 | 69 | 27 | 69 | 0.71 | 2.71 |
| Not always | 18 | 6,352 | 4 | 3 | 2 | (0.21, 2.33) | (0.66, 11.03) |
| **Regular hand hygiene** |  |  |  |  |  |  |  |
| Always | 374 | 81,247 | 71 | 27 | 71 | 0.72 | 5.68 |
| Not always | 15 | 6,499 | 3 | 3 | 1 | (0.22, 2.37) | (0.79, 40.87) |
| **Altered dining to decrease numbers or increase physical distancing** |  |  |  |  |  |  |  |
| Always | 257 | 65,365 | 60 | 23 | 60 | 0.24 ** | 1.57 |
| Not always | 24 | 3,411 | 4 | 5 | 2 | (0.09, 0.63) | (0.38, 6.40) |
| **Changed or increased bathroom facilities** |  |  |  |  |  |  |  |
| Always | 160 | 39,463 | 31 | 7 | 31 | 0.34 ** | 0.90 |
| Not always | 139 | 40,301 | 19 | 21 | 35 | (0.15, 0.80) | (0.56, 1.47) |

<sup>a</sup> Does not include camps not reporting data for examined NPI. <sup>b</sup> No. campers estimated from unique no. onsite campers.
<sup>c</sup> RR for always vs. not always. RRs for staffers determined using no. campers as denominator. \*\* p-value<0.05 \*\* p-value<0.00

For the 486 on-site camps serving 90,000 campers, the total number of confirmed cases equaled 30 and 72 cases, respectively, with 111 suspected and confirmed cases for campers and 191 suspected and confirmed cases reported for staff. A total of 74 camps reported having at least one confirmed COVID-19 case and 127 camps reported having at least one confirmed or suspected COVID-19 case. Of the camps with a confirmed case, 10 camps were overnight, 52 were day and 12 were combination overnight/day/rental camps. Five camps experienced more than five total cases among campers and staff, of which three experienced a COVID outbreak (>3 cases in a week). In the largest of these outbreaks, Camp A experienced 26 confirmed cases (9 camper, 17 staff) out of 66 overnight campers and 20 staff in one week. The NPIs instituted at Camp A were limited to regular hand sanitizing, increased cleaning, and pre-camp quarantining at home for campers and at home and camp for staff. The other two outbreaks occurred in day camps that used a wider range of NPIs. In Camp B, among a group of approximately 100 day campers and 30 staff, four confirmed cases in campers and three in staff during a one-week period were reported. This camp reported always requiring quarantining before camp, staff facial coverings, increasing cleaning, regular hand hygiene, and instituting measures to decrease its capacity and visitor access. Further, Camp B reported always or often instituting measures to increase physical distancing, through for example use of cohorts or pods or modified program, dining, and bathroom arrangements. However, camper facial coverings were not typical at Camp B. In the second day camp, Camp C, which had 199 day campers and 82 staff, four confirmed and nine suspected cases amongst staff were experienced during one week. At this camp, camper and staff facial coverings, physical distancing, cleaning, decreased visitors and decreased capacity, but not quarantine measures, were always or often in place.

For single site camps, the number of confirmed COVID-19 cases each week was low, absent the 3 observed outbreaks (Figure 1). Camper and staff COVID-19 cases occurred primarily in mid-summer, corresponding to weekly trends in the number of campers and staff and the overall US case rates. The higher number of confirmed cases in mid-summer in the South and West reflected community rates at that time, while those in the Northeast and Midwest largely reflected the three COVID-19 outbreaks.

Case rates were higher amongst campers in day as compared to overnight camps and camps with multiple program types, but the opposite pattern was observed for staff, with case rates for staff higher in overnight as compared to other camps. The incidence rate for COVID-19 cases amongst campers and staff was 3.3 and 8.03 per 10,000 campers, respectively. Incidence rates for cases amongst campers in overnight and day camps equaled 6.78 and 6.20, respectively, with a statistically insignificant relative risk (RR) of 1.09 (CI 95%: 0.51, 2.35), consistent with no difference in COVID-19 risks. For staff, RRs comparing overnight to day camps were also statistically insignificant at the 0.05 level, although the RR was larger than 1 (1.65, CI 95%: 0.99, 2.75) reflecting the outbreak in staff at one overnight camp. Across all on-site camps, risks of COVID-19 cases for campers and staff together were similar irrespective of whether the campers traveled from out-of-state to attend camp, with a statistically insignificant RR of 0.92 (95% CI: 0.61, 1.41).

The risk of COVID-19 cases was significantly reduced when campers or staff always wore facial coverings (Table 2). COVID-19 risks for campers were 0.36 (95% CI: 0.14, 0.95) and 0.39 (95% CI: 0.08, 0.40) times as high in camps with strict face covering policies for campers and staff, respectively, as compared to camps without, indicating that campers attending camps where face coverings were always worn had an approximately two-thirds reduction in risk of COVID-19 as compared to other campers. Correspondingly, COVID-19 risks were also reduced for staff working in camps where campers or staff always wore face coverings as compared to where they did not, with an 83% (RR: 0.17, 95% CI: 0.08, 0.40) and 62% (RR: 0.38, 95% CI: 0.24, 0.60) reduction in risks, respectively.

Pre-camp home quarantine measures for campers or staff were not found to significantly reduce risk of COVID-19 cases for either group. While no confirmed COVID-19 cases were experienced in camps requiring campers to quarantine at camp, the number of camps with such requirements was low, making their generalizability to other settings unclear.

Targeted physical distancing measures, including use of cohorts, pods, or bubbles, physical distancing, and modified programs to allow for physical distancing, were associated with less risk of COVID-19 cases in campers and to a lesser extent in staff. When evaluated individually, each of these physical distancing measures when always used produced similar reductions in risk, with rate reductions in campers of 61% (RR: 0.39, 95% CI: 0.19, 0.82) for cohorts, pods or bubbles, 69% (RR: 31, 95% CI 0.15, 0.65) for physical distancing measures, and 70% (RR: 0.30; 95% CI: 0.15, 0.61) for programs modified to increase physical distance. While statistically significant, risk reduction for staff was less pronounced when programs were modified for physical distancing, with ∼50% (RR: 0.52, 95% CI: 0.32, 0.82) reduction in COVID-19 risk as compared to staff working at other camps. While RRs were below 1, risks did not differ significantly for staff working at camps where cohorts or physical distancing NPIs were always as compared to not always in place. Risks for campers and staff were not reduced in camps that always decreased its capacity (campers: 1.44, 95% CI: 0.59, 3.52); staff: 3.77, 9 5% CI: 1.63, 8.72), suggesting that physical distancing behavior is needed for risk reduction rather than simply reduced numbers.

Since NPIs are often implemented together, we also examined RRs for specific pairwise NPI combinations (Table 3). As with individual NPIs, we found that constant camper facial coverings consistently lowered risks for campers and staff, regardless of what other NPI was also used. RRs were lowest in camps that always had both campers and staff wear facial coverings, with a 73% (RR: 0.27; 95% CI: 0.10, 0.73) and 87% (RR: 0.13; 95% CI: 0.06, 0.31) reduction in risks, respectively, as compared to camps where neither campers nor staff always wore facial coverings. This reduction in risks were greater than that when only staff, but not campers, wore facial coverings, suggesting the greater importance of camper facial coverings as an NPI.

**Table 3.** Effect of Multiple NPI Combinations on Total (Camper and Staff) COVID-19 Cases^a^

| NPI | Response | Number |  | Number of Cases |  |  | RR (95% CI) |  |
| --- | --- | --- | --- | --- | --- | --- | --- | --- |
|  |  | Camps | Campers | Camps | Camper | Staff | Camper | Staff |
| Camper and Staff Facial Covering | Neither | 118 | 26,589 | 18 | 16 | 39 | - | - |
|  | Camper Facial Covering Only | 0 | 0 | 0 | 0 | 0 | - | - |
|  | Staff Facial Covering Only | 137 | 29,587 | 41 | 9 | 24 | 0.51 (0.22, 1.14) | 0.55 (0.33, 0.92) |
|  | Both | 126 | 30,883 | 13 | 5 | 6 | 0.27 (0.10, 0.73) | 0.13 (0.06, 0.31) |
| Camper Facial Covering and Home Camper Quarantine | Neither | 195 | 42,087 | 47 | 11 | 29 | - | - |
|  | Camper Facial Covering Only | 87 | 17,547 | 6 | 2 | 2 | 0.44 (0.10, 1.97) | 0.17 (0.04, 0.69) |
|  | Home Camper Quarantine Only | 56 | 14,029 | 12 | 14 | 34 | 3.82 (1.73, 8.41) | 3.52 (2.14, 5.77) |
|  | Both | 34 | 10,786 | 4 | 2 | 4 | 0.71 (0.16, 3.20) | 0.54 (0.19, 1.53) |
| Camper Facial Covering and Physical Distancing | Neither | 110 | 25,477 | 17 | 18 | 32 | - | - |
|  | Camper Facial Covering Only | 21 | 5,019 | 1 | 1 | 0 | 0.28 (0.04, 2.11) | 0.00 (0.00, NA) |
|  | Physical Distancing Only | 145 | 30,699 | 42 | 7 | 31 | 0.32 (0.13, 0.77) | 0.80 (0.49, 1.32) |
|  | Both | 105 | 25,864 | 12 | 4 | 6 | 0.22 (0.07, 0.65) | 0.18 (0.08, 0.44) |
| Camper Facial Coverings and Modified Programs | Neither | 85 | 23,113 | 15 | 17 | 31 | - | - |
|  | Camper Facial Coverings Only | 4 | 662 | 0 | 0 | 0 | 0.00 (0.00, NA) | 0.00 (0.00, NA) |
|  | Modified Programs Only | 167 | 31,384 | 44 | 8 | 32 | 0.35 (0.15, 0.80) | 0.76 (0.46, 1.25) |
|  | Both | 120 | 29,285 | 13 | 5 | 6 | 0.23 (0.09, 0.63) | 0.15 (0.06, 0.37) |
| Physical Distancing and Modified Programs | Neither | 79 | 19,504 | 14 | 17 | 30 | - | - |
|  | Physical Distancing Only | 12 | 4,391 | 1 | 0 | 1 | 0.00 (0.00, NA) | 0.15 (0.02, 1.09) |
|  | Modified Programs Only | 54 | 11,112 | 4 | 2 | 2 | 0.21 (0.05, 0.89) | 0.12 (0.03, 0.49) |
|  | Both | 238 | 50,177 | 55 | 11 | 39 | 0.25 (0.12, 0.54) | 0.51 (0.31, 0.81) |
| Physical Distancing and Home Camper Quarantine | Neither | 99 | 21,689 | 10 | 4 | 8 | - | - |
|  | Physical Distancing Only | 186 | 38,565 | 44 | 9 | 24 | 1.27 (0.39, 4.11) | 1.69 (0.76, 3.75) |
|  | Home Camper Quarantine Only | 32 | 8,807 | 8 | 15 | 24 | 9.24 (3.07, 27.82) | 7.39 (3.32, 16.44) |
|  | Both | 61 | 16,008 | 9 | 1 | 16 | 0.34 (0.04, 3.03) | 2.71 (1.16, 6.33) |
| Modified Programs and Home Camper Quarantine | Neither | 70 | 16,915 | 10 | 3 | 9 | - | - |
|  | Modified Programs Only | 212 | 40,724 | 44 | 10 | 23 | 1.38 (0.38, 5.03) | 1.06 (0.49, 2.29) |
|  | Home Camper Quarantine Only | 19 | 6,860 | 5 | 14 | 22 | 11.51 (3.31, 40.03) | 6.03 (2.78, 13.08) |
|  | Both | 72 | 17,955 | 12 | 2 | 18 | 0.63 (0.10, 3.76) | 1.88 (0.85, 4.19) |

<sup>a</sup> Comparisons made using subset of data for which there is complete data for each examined NPI. RRs calculated comparing 1 or 2 NPIs to neither NPI.

We found an approximate 80% reduction in risks for both campers (RR: 0.22; 95% CI: 0.07, 0.65) and staff (RR: 0.18; 95% CI: 0.08, 0.44), when camps always implemented both camper facial coverings and a physical distancing NPI, as compared to when neither was always implemented. This reduction in risks was larger than that when only physical distancing NPIs were always implemented. When two targeted physical distancing NPIs – physical distancing and modified programs to increase physical distancing – were always implemented together, RRs were significantly lower as compared to camps where neither were always implemented. However, risks for camps implementing both physical distancing NPIs were similar to camps implementing just one physical distancing NPI. The additional use of pre-camp home quarantines did not reduce risks beyond that afforded by physical distancing or facial coverings alone.

## Discussion

In our sample of 486 camps operating on-site during summer 2020, the number of COVID-19 cases was low, with an incidence rate for campers and staff of 3.35 and 8.03 per 10,000 campers, resulting in a total of 30 and 72 confirmed cases, respectively. COVID-19 case incidence rates were not statistically different for overnight and day camps. COVID-19 case rates in camps are low relative to overall rates in the US during the same period, which in June 1 were estimated to equal approximately 55 cases per 10,000 people, increasing to 184 cases per 10,000 people by August 31 [5]. A range of NPIs were instituted at the camps, with risk ratios for individual and multiple NPIs consistently demonstrating the importance of constant facial coverings, especially by campers, and of targeted physical distancing measures.

Our findings are consistent with evidence showing lower incidence rates in children overall [6] and within congregate settings [7], reduced risks of SARS-CoV-2 infection with the use of facial coverings and physical distancing [2,8,9,10] and with anecdotal evidence of SARS-CoV-2 outbreaks in camps where camper and often staff face coverings was not universal [3,4,11]. For example, in a study of COVID-19 cases and transmission in 17 Wisconsin schools, authors reported low transmission rates in schools that required student masking and cohorting [10]. Correspondingly, In a study of almost 2 million confirmed COVID-19 cases from 190 countries [12], for example, the individual and combined effectiveness of mandatory public face coverings, quarantine, social distancing, and traffic restrictions on the COVID-19 effective reproduction number (R_t_) was examined. The study showed the greatest reductions in R_t_ for physical distancing measures and the simultaneous implementation of 2+ NPIs, with individual implementation of any of the examined NPIs, including mandatory facial coverings, also reducing R_t_ of COVID-19. As in this study, we also found physical distancing measures to effectively reduce risks. However, we found camper facial coverings to offer greater risk reduction, likely reflective of (1) our study being within a child congregate setting, where children and staff spend considerable time together and physical distancing measures are harder to enforce and (2) the ability of camper masks when always worn to reduce risks of SARS-CoV-2 transmission during the period before a COVID-19 case can be identified.

NPIs related to decreased capacity and pre-camp quarantining were not found by themselves to be significant COVID-19 risk reduction measures, which particularly in the case of pre-camp quarantining was surprising given evidence of its effectiveness in other settings. For example, successful operation of four Maine overnight camps during Summer 2020 was attributed in part to implementation of strict prearrival quarantines and pre- and post-arrival testing, which identified asymptomatic COVID-19 cases prior to the start of camp, and with subsequent isolation and contact quarantining, prevented further SARS-CoV-2 transmission [2]. These camps also instituted face coverings, targeted physical distancing measures, and other NPIs. Our results suggest that face coverings and targeted physical distancing measures in particular were important contributors to the lack of COVID-19 cases at the Maine camps.

Our study has several limitations. First, our findings are based on aggregate data reported by camps, and as such, measures of NPI usage and COVID-19 cases were not independently verified. As a result, COVID-19 cases were likely underestimated due to under-reporting and lack of detection of asymptomatic cases. It is also possible that NPI usage was underestimated or overestimated. Second, our findings may be affected by selection bias, resulting from the self-selection of camps voluntarily participating in our survey. Demographics of camps participating in the survey mirrored that of the overall camp population, suggesting that selection bias was minimal. Third, completion rates of several survey questions were below 75%, suggesting the potential for reporting bias in regards to what information they chose to report.

These limitations are balanced by our substantial study strengths, including our nationwide cohort of day and overnight camps and the associated data documenting COVID-19 cases and the usage and adherence to numerous NPIs. These data allowed us to characterize COVID-19 risks to both campers and staff and to assess the effectiveness of NPIs, including facial coverings, physical distancing, quarantine, and cleaning measures. Our findings show rates of COVID-19 cases for campers and staff were relatively low – even in areas with high community COVID-19 rates – and further demonstrate the importance of strict face covering and targeted physical distancing measures to reduce SARS-CoV-2 infection in campers and staff.

## Data Availability

Data are available upon request.

## Abbreviations

COVID-19: coronavirus disease 2019
NPI: non-pharmaceutical interventions

